# Diagnostic Concordance of Immediate Versus 1-Hour Technetium-99m Hydroxymethylene Diphosphonate Scintigraphy in Suspected Transthyretin Amyloid Cardiomyopathy

**DOI:** 10.64898/2026.06.16.26355752

**Authors:** Andrew Costa, Austen Suits, Jack Miller, Maros Ferencik, David M. German, Evan F. Shalen, Gagandeep Choudhary, Laszlo Szidonya, Mike Nguyen, Nadine Mallak, Sebastian Obrzut, Ahmad Masri

**Affiliations:** Department of Internal Medicine, Oregon Health and Science University, Portland, OR, USA; Oregon Health and Science University College of Medicine, Portland, OR, USA; Knight Cardiovascular Institute, Oregon Health and Science University, Portland, OR, USA; Department of Diagnostic Radiology, Oregon Health & Science University, Portland, OR, USA

**Author notes:** Corresponding author: Ahmad Masri, MD, MS, Associate Professor of Medicine, Mail code: UHN-62, 3181 SW Sam Jackson Rd, Portland, OR 97239.

**Keywords:** Transthyretin Amyloid Cardiomyopathy (ATTR-CM), Technetium-99m Hydroxymethylene Diphosphonate (^99m^Tc-HDP), Single-Photon Emission Computed Tomography / Computed Tomography (SPECT/CT), Target to Background Ratio (TBR), Perugini

## Abstract

**Background:** Bone-avid tracer myocardial scintigraphy for the diagnosis of transthyretin amyloid cardiomyopathy (ATTR-CM) has traditionally employed imaging at one or 3-hour intervals. Technetium-99m hydroxymethylene diphosphonate (^99m^Tc-HDP) has unique characteristics that may enable earlier imaging. We investigated the diagnostic concordance of immediate versus 1-hour acquisitions.

**Methods:** Consecutive patients with suspected ATTR-CM underwent planar imaging and SPECT/CT immediately and at 1-hour following the administration of ^99m^Tc-HDP. Perugini grades and heart to contralateral lung (H/CL) ratios were assessed. Target-to-background ratios (TBRs) were calculated on the SPECT/CT acquisitions using the left ventricular (LV) septum and three background regions: aorta, LV blood-pool, and vertebrae. We assessed diagnostic concordance using Cohen’s Kappa (*κ*), temporal stability using paired *t*-tests, and correlation between timepoints using Pearson’s coefficient (*r*). The 1-hour SPECT/CT interpretation served as the protocol reference standard.

**Results:** Forty-eight patients (83% male; median age, 80 [73–85] years) were evaluated. One-hour SPECT/CT identified 19 positive and 29 negative cases. Immediate SPECT/CT demonstrated 100% diagnostic concordance with the 1-hour reference standard (*κ* = 1.000; *p* < 0.001). The LV septum/LV Blood-Pool TBR showed the highest correlation (*r* = 0.956; *p* < 0.001). The LV Septum/Aorta TBR demonstrated high correlation *(r* = 0.918; *p* < 0.001) and remained stable in the ATTR-negative cohort (−0.02; *p* = 0.54). Significant decreases in the LV Septum/Vertebrae TBR were observed in the ATTR-negative (−0.55; *p* < 0.001) and ATTR-positive cohorts (−1.14; *p* < 0.001).

**Conclusions:** In this cohort of patients with suspected ATTR-CM, immediate ^99m^Tc-HDP SPECT/CT demonstrated high diagnostic concordance with standard 1-hour acquisitions. This expedited approach may improve nuclear laboratory throughput and patient satisfaction.

## Introduction

While once considered to be a rare clinical entity, transthyretin amyloid cardiomyopathy (ATTR-CM) is an increasingly recognized cause of heart failure that is associated with high rates of morbidity and mortality in older adults [1]. To address the diagnostic demand created by increased clinical recognition of its prevalence, noninvasive bone-avid tracer myocardial scintigraphy has emerged as an integral part of the evaluation of ATTR-CM [2]. Bone-avid tracer myocardial scintigraphy protocols have traditionally employed imaging at one or 3-hour intervals to allow for sufficient blood-pool clearance to maximize diagnostic accuracy; however, these conventional intervals may not be optimal when evaluating tracer performance across earlier imaging timepoints [3,4]. Recently, attempts to refine imaging protocols by minimizing the delay between tracer administration and image acquisition have sought to alleviate the burden of prolonged wait times for an elderly patient population while simultaneously enhancing laboratory efficiency [4,5,6,7].

Due to recurrent shortages of technetium-99m pyrophosphate (^99m^Tc-PYP), technetium-99m hydroxymethylene diphosphonate (^99m^Tc-HDP) has increasingly been utilized in clinical practice and possesses unique kinetic characteristics that may enable even earlier imaging [4,8,9]. While other investigators have explored accelerated planar scintigraphy with ^99m^Tc-HDP, the two-dimensional nature of planar imaging is inherently limited by blood-pool interference and soft tissue overlap [10,11]. In recent years, single-photon emission computed tomography in conjunction with computed tomography (SPECT/CT) has been instrumental to mitigate these limitations [3]. Recent investigations have shown that 1-hour ^99m^Tc-HDP SPECT/CT is feasible with excellent diagnostic capabilities [5,6].

Despite continued interest in accelerated protocols, the diagnostic performance of static, immediate-phase imaging—the most widely accessible and reproducible approach for traditional nuclear medicine laboratories—utilizing ^99m^Tc-HDP remains uncharacterized. We hypothesized that ^99m^Tc-HDP would be a suitable candidate for an accelerated imaging protocol given its advantageous pharmacokinetic profile characterized by rapid blood-pool clearance driven by efficient skeletal uptake [4,8,9,12]. The objective of this study was to systematically evaluate the diagnostic concordance of immediate-phase versus 1-hour static ^99m^Tc-HDP SPECT/CT acquisitions in cases of suspected ATTR-CM.

## Methods

### Study Design and Patient Population

We evaluated a cohort of patients who underwent a quality-related imaging intervention to assess the clinical performance of ^99m^Tc-HDP during national shortage of ^99m^Tc-PYP. Patients with suspected ATTR-CM presenting for a clinically indicated ^99m^Tc-HDP scan were included in the imaging protocol. The primary objective of this study was to assess the quality of and evaluate the diagnostic concordance between immediate-phase and standard 1-hour ^99m^Tc-HDP SPECT acquisitions. Diagnostic concordance was defined as binary visual agreement on SPECT/CT, using a Perugini-like scale (grade 0–1 vs grade 2– 3) between immediate and standard 1-hour acquisitions. Imaging was performed prospectively per protocol; baseline clinical and laboratory data were collected retrospectively from the medical record. Patients with suspected cardiac amyloidosis were enrolled in a registry approved by the OHSU Institutional Review Board.

### Static, Dual-Phase Image Acquisition Protocol and Image Reconstruction

All patients were imaged on either a Siemens Symbia Intevo Bold or a Siemens Symbia Pro.specta X3 dual-head nuclear gamma camera, both equipped with integrated CT and low-energy high-resolution (LEHR) collimators. A mean (± standard deviation) activity of 14.7 ± 1.0 millicuries (544 ± 35 MBq) of ^99m^Tc-HDP was administered prior to imaging. Immediately following the administration of ^99m^Tc-HDP, an anterior-posterior planar image was acquired, which served as time zero (*T*_0_) for each patient across the protocol. Subsequently, SPECT/CT was acquired, and at 1-hour after *T*_0_, repeat planar and SPECT images were performed. For both immediate and 1-hour planar acquisitions, 750,000 counts were implemented using a 256×256 matrix and a zoom of 1.00. Images were obtained using a 140-keV photopeak with a ± 10% energy window. After planar acquisitions, three-dimensional SPECT was performed in step-mode using a 128×128 matrix to ensure temporal consistency between the planar and SPECT images. The acquisition included 64 views at 15 seconds per view with a 180-degree detector configuration and a non-circular orbit to minimize the detector-to-patient distance. A low-dose CT was performed for the purposes of attenuation correction and anatomical localization. SPECT data were reconstructed using an Ordered Subset Expectation Maximization (OSEM) algorithm (10 iterations, 8 subsets) on AutoRecon software. Attenuation and scatter correction were integrated into the reconstruction process using maps derived from a low-dose CT scan. A 9 mm Gaussian post-reconstruction filter was then applied to the corrected SPECT data. The CT field of view was standardized to include the entire thorax to ensure consistent coverage of the myocardium and reference structures.

### Image Interpretation and Reference Standards

Perugini semiquantitative evaluation (grade 0–1 vs grade 2–3) and quantitative heart to contralateral lung (H/CL) ratios were assessed on planar images at both timepoints. A Perugini scale (grade 0–1 vs grade 2– 3) was also applied to SPECT/CT images. Cross-sectional SPECT/CT resolution enabled direct comparison of osseous tracer uptake of the ribs, sternum, and vertebrae to myocardium. Perugini grading ≥ 2 with an H/CL ratio ≥ 1.5 and SPECT Perugini grading ≥ 2 in conjunction with diffuse myocardial uptake were considered positive for the planar and SPECT/CT acquisitions, respectively. The protocol reference standard was the 1-hour SPECT/CT as interpreted by an experienced nuclear cardiologist and a nuclear medicine radiologist. Immediate-phase planar and SPECT/CT acquisitions were then independently interpreted by a separate reader who was blinded to the results of the 1-hour SPECT/CT reference standard. Blinded inter-observer and intra-observer variability was assessed for 10% of randomly selected patients at both imaging timepoints for the two imaging modalities. A two-month interval separated the initial and repeat grading sessions for intra-observer variability analysis.

### Quantitative Image Analysis and Contour Placement

Quantitative analysis utilizing regions/volumes of interest was performed for both the planar and SPECT/CT acquisitions to objectively characterize ^99m^Tc-HDP tissue distribution and myocardial uptake between timepoints. For each planar acquisition, a 7.05 ± 0.13cm^2^ circular region of interest (ROI) was placed over the cardiac silhouette. To maintain consistency across ROIs, the initial ROI was duplicated and mirrored over the contralateral chest wall. The H/CL ratio was derived from the mean counts of the mirrored ROIs. For the SPECT/CT acquisitions, volumes of interest (VOIs) were drawn over four regions: LV (left ventricular) septum, vertebra, LV blood-pool, and descending aorta. Mean counts were recorded for each VOI at both timepoints. To ensure reproducibility and anatomical correspondence between timepoints, a consistent cranio-caudal CT level was maintained for each patient when defining VOIs across all four references. Target-to-background ratios (TBRs) using mean counts obtained from the VOIs were calculated to compare LV septum signal intensity relative to the vertebrae, descending aorta, and LV blood-pool. MIM imaging software (Version 7.4.2) was used for all qualitative and quantitative imaging analysis.

### Statistical Analysis

Categorical variables were presented as frequencies and percentages. Because data were collected retrospectively, comorbidities not documented in the medical record were classified as absent, and percentages for these variables were calculated using all patients in each cohort. For laboratory values, the number of patients with available data is reported in Table 1, and values below the assay detection limit were set to the limit value. Differences between cohorts for categorical data were assessed using Fisher’s exact test, with the Fisher-Freeman-Halton exact test used for variables with more than two categories (race and NYHA class). Baseline clinical and demographic variables and imaging acquisition intervals demonstrated non-normal distributions and were expressed as medians with interquartile ranges (IQR). Differences between cohorts for these variables were assessed using the non-parametric Mann-Whitney U test. Quantitative imaging metrics (H/CL ratios and TBRs) are expressed as means ± SD. Differences in imaging metrics between the ATTR-positive and ATTR-negative cohorts were evaluated using independent *t*-tests. Paired differences between immediate and 1-hour acquisitions within the same cohort satisfied the normality assumption on Shapiro-Wilk testing (all *p* > 0.05) and were assessed using paired *t*-tests. The diagnostic performance of the immediate SPECT/CT and planar imaging was evaluated by calculating sensitivity and specificity compared to the 1-hour SPECT/CT reference standard. Corresponding 95% confidence intervals (CI) for diagnostic proportions were calculated using the Wilson score interval method. Binary diagnostic concordance (negative: Perugini 0-1 vs positive: Perugini 2-3) was quantified using Cohen’s kappa (*κ*) coefficient. Linear association of quantitative metrics was assessed using Pearson’s correlation coefficient (*r*). 95% CIs for Pearson’s *r* were calculated using the Fisher z transformation. To account for potential spectrum bias arising from bimodal cohort separation, correlation analyses were performed across the pooled population and stratified within the ATTR-positive and ATTR-negative cohorts independently. Agreement between the immediate and 1-hour quantitative measurements was further evaluated using Bland-Altman analysis. Results are reported as the point estimate for the mean bias along with its 95% CI and the 95% limits of agreement (LoA) to assess both systematic bias and individual variability. Reliability of the immediate-phase interpretations was established through blinded inter-observer and intra-observer variability analyses of 10% of cases that were randomly selected. A two-tailed *p* value < 0.05 was defined as the threshold for statistical significance. Sample size was estimated to ensure adequate power to detect diagnostic concordance between the immediate and 1-hour protocols. Assuming a sensitivity of 95% and specificity of 90% for the 1-hour protocol relative to established non-invasive diagnostic criteria for ATTR-CM [3], a minimum of 39 subjects (approximately 35% with positive scans) was estimated to provide 80% power to detect the anticipated level of agreement using a paired-sample design. All statistical analyses were conducted using Python (Version 3.11).

**Table 1.**
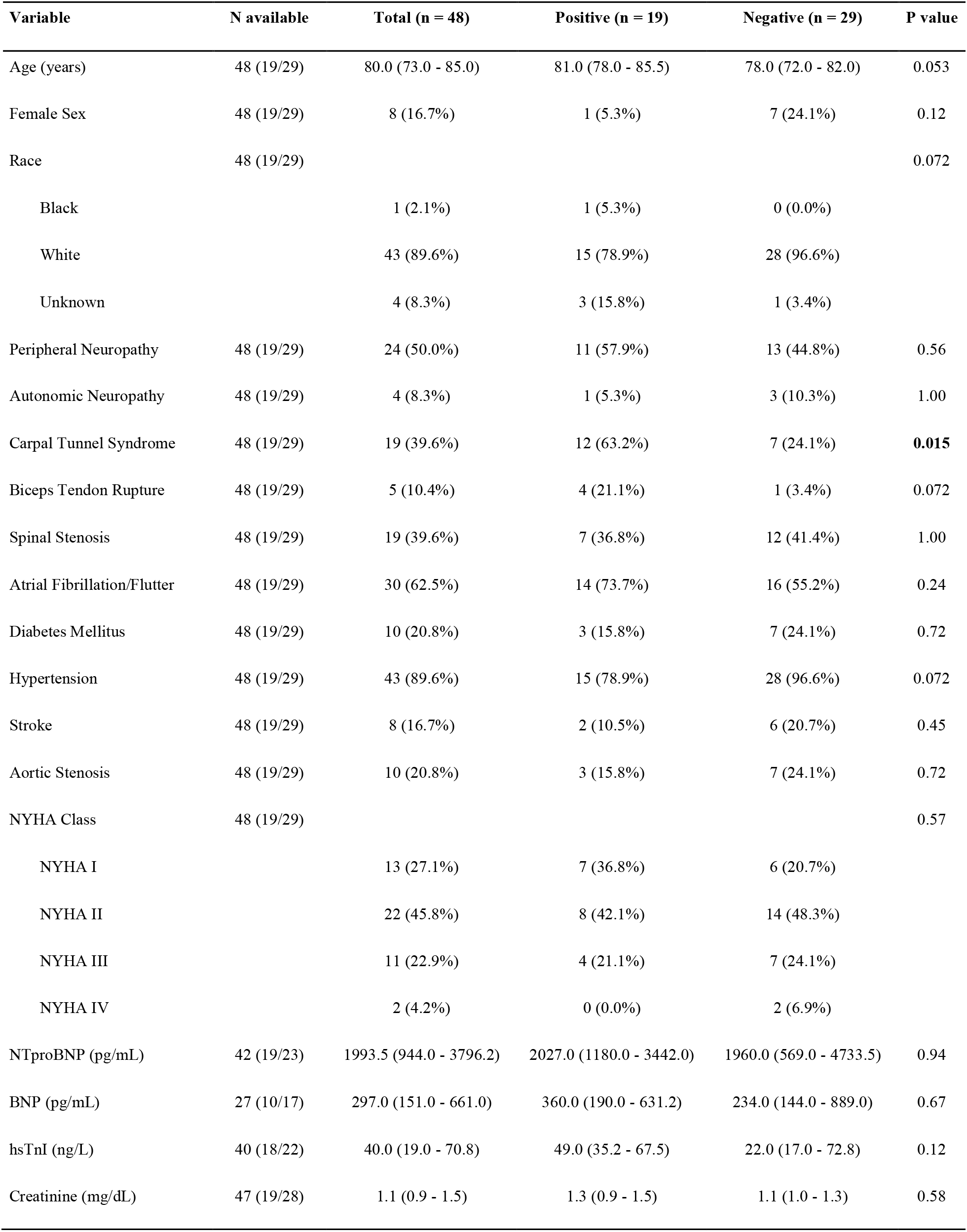
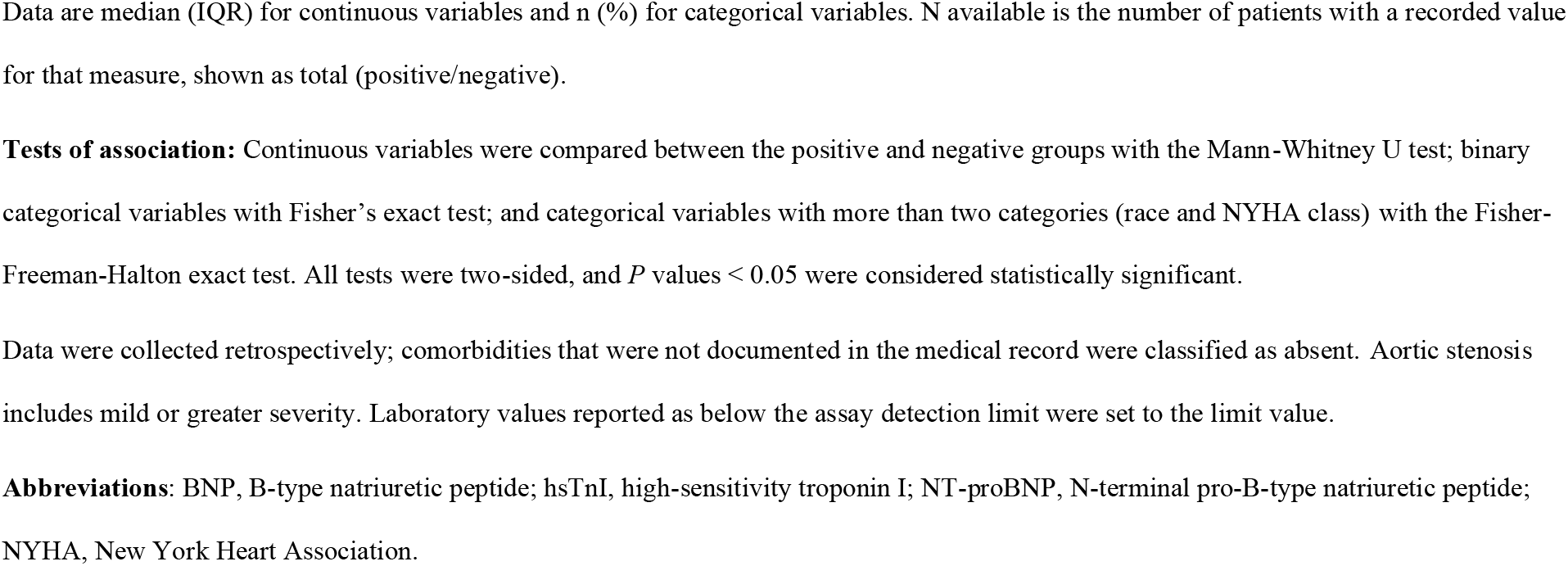
Baseline demographics and clinical characteristics of the study cohort.

| Variable | N available | Total (n = 48) | Positive (n = 19) | Negative (n = 29) | P value |
| --- | --- | --- | --- | --- | --- |
| Age (years) | 48 (19/29) | 80.0 (73.0 - 85.0) | 81.0 (78.0 - 85.5) | 78.0 (72.0 - 82.0) | 0.053 |
| Female Sex | 48 (19/29) | 8 (16.7%) | 1 (5.3%) | 7 (24.1%) | 0.12 |
| Race | 48 (19/29) |  |  |  | 0.072 |
| Black |  | 1 (2.1%) | 1 (5.3%) | 0 (0.0%) |  |
| White |  | 43 (89.6%) | 15 (78.9%) | 28 (96.6%) |  |
| Unknown |  | 4 (8.3%) | 3 (15.8%) | 1 (3.4%) |  |
| Peripheral Neuropathy | 48 (19/29) | 24 (50.0%) | 11 (57.9%) | 13 (44.8%) | 0.56 |
| Autonomic Neuropathy | 48 (19/29) | 4 (8.3%) | 1 (5.3%) | 3 (10.3%) | 1.00 |
| Carpal Tunnel Syndrome | 48 (19/29) | 19 (39.6%) | 12 (63.2%) | 7 (24.1%) | **0.015** |
| Biceps Tendon Rupture | 48 (19/29) | 5 (10.4%) | 4 (21.1%) | 1 (3.4%) | 0.072 |
| Spinal Stenosis | 48 (19/29) | 19 (39.6%) | 7 (36.8%) | 12 (41.4%) | 1.00 |
| Atrial Fibrillation/Flutter | 48 (19/29) | 30 (62.5%) | 14 (73.7%) | 16 (55.2%) | 0.24 |
| Diabetes Mellitus | 48 (19/29) | 10 (20.8%) | 3 (15.8%) | 7 (24.1%) | 0.72 |
| Hypertension | 48 (19/29) | 43 (89.6%) | 15 (78.9%) | 28 (96.6%) | 0.072 |
| Stroke | 48 (19/29) | 8 (16.7%) | 2 (10.5%) | 6 (20.7%) | 0.45 |
| Aortic Stenosis | 48 (19/29) | 10 (20.8%) | 3 (15.8%) | 7 (24.1%) | 0.72 |
| NYHA Class | 48 (19/29) |  |  |  | 0.57 |
| NYHA I |  | 13 (27.1%) | 7 (36.8%) | 6 (20.7%) |  |
| NYHA II |  | 22 (45.8%) | 8 (42.1%) | 14 (48.3%) |  |
| NYHA III |  | 11 (22.9%) | 4 (21.1%) | 7 (24.1%) |  |
| NYHA IV |  | 2 (4.2%) | 0 (0.0%) | 2 (6.9%) |  |
| NTproBNP (pg/mL) | 42 (19/23) | 1993.5 (944.0 - 3796.2) | 2027.0 (1180.0 - 3442.0) | 1960.0 (569.0 - 4733.5) | 0.94 |
| BNP (pg/mL) | 27 (10/17) | 297.0 (151.0 - 661.0) | 360.0 (190.0 - 631.2) | 234.0 (144.0 - 889.0) | 0.67 |
| hsTnI (ng/L) | 40 (18/22) | 40.0 (19.0 - 70.8) | 49.0 (35.2 - 67.5) | 22.0 (17.0 - 72.8) | 0.12 |
| Creatinine (mg/dL) | 47 (19/28) | 1.1 (0.9 - 1.5) | 1.3 (0.9 - 1.5) | 1.1 (1.0 - 1.3) | 0.58 |

Data are median (IQR) for continuous variables and n (%) for categorical variables. N available is the number of patients with a recorded value for that measure, shown as total (positive/negative).
**Tests of association:** Continuous variables were compared between the positive and negative groups with the Mann-Whitney U test; binary categorical variables with Fisher's exact test; and categorical variables with more than two categories (race and NYHA class) with the Fisher-Freeman-Halton exact test. All tests were two-sided, and *P* values < 0.05 were considered statistically significant.
Data were collected retrospectively; comorbidities that were not documented in the medical record were classified as absent. Aortic stenosis includes mild or greater severity. Laboratory values reported as below the assay detection limit were set to the limit value.
**Abbreviations:** BNP, B-type natriuretic peptide; hsTnI, high-sensitivity troponin I; NT-proBNP, N-terminal pro-B-type natriuretic peptide; NYHA, New York Heart Association.

## Results

### Cohort Demographics and Clinical Characteristics

A total of 48 patients with suspected ATTR-CM between July and November 2024 completed the imaging protocol. Of those, 19 (39.6%) individuals had positive and 29 (60.4%) had negative ^99m^Tc-HDP SPECT/CT acquisitions at 1-hour. Overall, clinical characteristics and demographics were comparable between the two cohorts (all *p* ≥ 0.05), with the exception of carpal tunnel syndrome (ATTR-positive: 63.2% vs ATTR-negative 24.1%; *p* = 0.015), Table 1. Patients were reflective of a typical amyloid referral population, with a median age of 80.0 years (IQR 73–85); 83.3% were male. Patients also had a high prevalence of clinical findings associated with amyloidosis, including atrial fibrillation/flutter (62.5%) and spinal stenosis (39.6%). Most patients were symptomatic with 72.9% exhibiting New York Heart Association (NYHA) Class II or higher heart failure symptoms. In the ATTR-negative cohort, one patient was diagnosed with AL amyloidosis after participation in the protocol. Zero patients in the ATTR-positive cohort were diagnosed with AL amyloidosis during follow-up.

### Imaging Acquisition Intervals

Following administration of 14.7 ± 1.0 mCi (544 ± 35 MBq) of ^99m^Tc-HDP, immediate-phase SPECT/CT images were acquired at a median of 3.8 (IQR 2.4-11.4) minutes after tracer administration. One-hour planar imaging and SPECT/CT occurred at a median of 54.5 (IQR 51.3-59.1) and 57.7 (IQR 55.0-63.5) minutes after tracer administration, respectively. No adverse events were observed from either the immediate-phase or 1-hour ^99m^Tc-HDP acquisitions.

### Immediate Planar Imaging and Quantitative Analysis

Immediate-phase planar imaging demonstrated high sensitivity of 94.7% (95% CI: 75.4% to 99.1%) but poor specificity of 31.0% (95% CI: 17.3% to 49.2%) in comparison to the 1-hour SPECT/CT reference standard. Diagnostic concordance between immediate planar and 1-hour SPECT/CT was poor (*κ* = 0.220; 95% CI: −0.03 to 0.47; *p* = 0.09). Full planar data can be found in the Planar Supplement section.

### Diagnostic Concordance of Immediate and 1-Hour SPECT/CT Visual Grading

Immediate-phase SPECT/CT acquisitions demonstrated 100% sensitivity (95% CI: 83.2% to 100%) and 100% specificity (95% CI: 88.3% to 100%) relative to the 1-hour reference standard. There was similar diagnostic concordance (48/48 patients) between the two timepoints (*κ* = 1.000; 95% CI: 1.00 to 1.00; *p* < 0.001), Figure 1.

**Figure 1.**
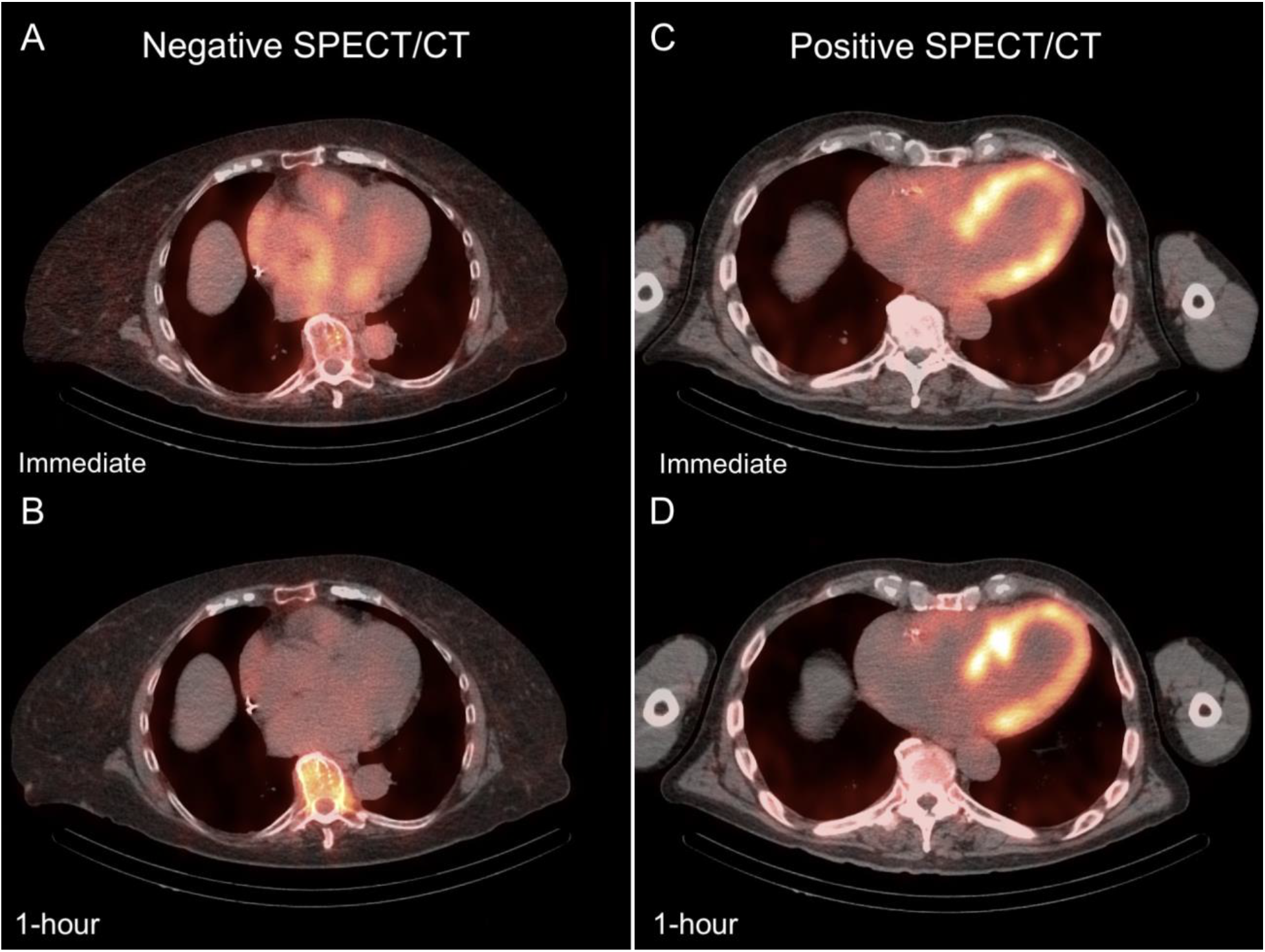
Comparison of immediate and 1-hour ^99m^Tc-HDP SPECT/CT acquisitions in a patient without ATTR-CM (negative scan, A: immediate, B: 1-hour) and a patient with ATTR-CM (positive scan, C: immediate, D: 1-hour) **Abbreviations:** ATTR-CM, Transthyretin amyloid cardiomyopathy; SPECT/CT, Single-photon emission computed tomography/computed tomography; ^99m^Tc-HDP, technetium-99m hydroxymethylene diphosphonate.

### Left Ventricular Blood-Pool Reference

These measures were highly correlated between timepoints, *r* = 0.956 (95% CI: 0.922 to 0.975; *p* < 0.001), Table 2 and Supplemental Figure 1A, and were distinct between cohorts during immediate-phase imaging (ATTR-negative: 0.70 ± 0.09 vs ATTR-positive: 2.01 ± 0.41; *p* < 0.001). There was a small increase in mean TBRs between timepoints in the ATTR-negative cohort (+0.06; 95% CI: 0.03 to 0.09; *p* < 0.001) and ATTR-positive cohort (+0.41; 95% CI: 0.21 to 0.62; *p* < 0.001), Table 3.

**Table 2.** Pooled and intra-cohort correlation between immediate and 1-hour ^99m^Tc-HDP SPECT/CT TBRs.

| TBR Reference Site | Pearson's r | 95% CI | P value |
| --- | --- | --- | --- |
| LV Septum/LV Blood-Pool |  |  |  |
| Pooled (n = 48) | 0.956 | 0.922 to 0.975 | <**0.001** |
| Negative (n = 29) | 0.720 | 0.480 to 0.860 | <**0.001** |
| Positive (n = 19) | 0.763 | 0.473 to 0.904 | <**0.001** |
| LV Septum/Aorta |  |  |  |
| Pooled (n = 48) | 0.918 | 0.857 to 0.953 | <**0.001** |
| Negative (n = 29) | 0.556 | 0.238 to 0.766 | **0.002** |
| Positive (n = 19) | 0.559 | 0.141 to 0.808 | **0.013** |
| LV Septum/Vertebrae |  |  |  |
| Pooled (n = 48) | 0.905 | 0.835 to 0.946 | <**0.001** |
| Negative (n = 29) | 0.228 | -0.151 to 0.549 | 0.234 |
| Positive (n = 19) | 0.517 | 0.082 to 0.787 | **0.023** |

**Abbreviations:** CI, confidence interval; r, Pearson's r; SPECT/CT, single-photon emission computed tomography/computed tomography; TBR, target-to-background ratio; $^{99m}\text{Tc}$ -HDP, technetium-99m hydroxymethylene diphosphonate.

**Table 3.** Analysis of TBR stability and agreement between immediate and 1-hour 99mTc-HDP SPECT/CT acquisitions.

| TBR Site | Cohort | Mean Bias | 95% CI | 95% LoA | P value |
| --- | --- | --- | --- | --- | --- |
| LV Septum/LV Blood-Pool | Negative | 0.06 | +0.03 to +0.09 | -0.10 to +0.22 | <**0.001** |
|  | Positive | 0.41 | +0.21 to +0.62 | -0.43 to +1.26 | <**0.001** |
| LV Septum/Aorta | Negative | -0.02 | -0.08 to +0.04 | -0.32 to +0.29 | 0.54 |
|  | Positive | 0.71 | +0.36 to +1.06 | -0.71 to +2.13 | <**0.001** |
| LV Septum/Vertebrae | Negative | -0.55 | -0.64 to -0.46 | -0.99 to -0.11 | <**0.001** |
|  | Positive | -1.14 | -1.39 to -0.89 | -2.17 to -0.11 | <**0.001** |

**Note:** Mean bias is defined as 1-hour value minus immediate value.
**Abbreviations:** CI, confidence interval; LoA, limits of agreement; SPECT/CT, single-photon emission computed tomography/computed tomography; TBR, target-to-background ratio; 99mTc-HDP, technetium-99m hydroxymethylene diphosphonate.

### Descending Aorta Reference

These measures were also highly correlated between timepoints, *r* = 0.918 (95% CI: 0.857 to 0.953; *p* < 0.001), Table 2 and Figure 2A, and successfully differentiated cohorts during immediate-phase imaging (ATTR-negative: 0.76 ± 0.16 vs ATTR-positive: 2.40 ± 0.60; *p* < 0.001). Notably, this site behaved as a stable reference in the negative cohort, where the ratio remained unchanged between timepoints (−0.02; 95% CI: −0.08 to 0.04; *p* = 0.54), Table 3 and Figure 3A. In the ATTR-positive cohort, the mean TBR increased significantly over time (+0.71; 95% CI: 0.36 to 1.06; *p* < 0.001), Table 3 and Figure 3A.

**Figure 2.**
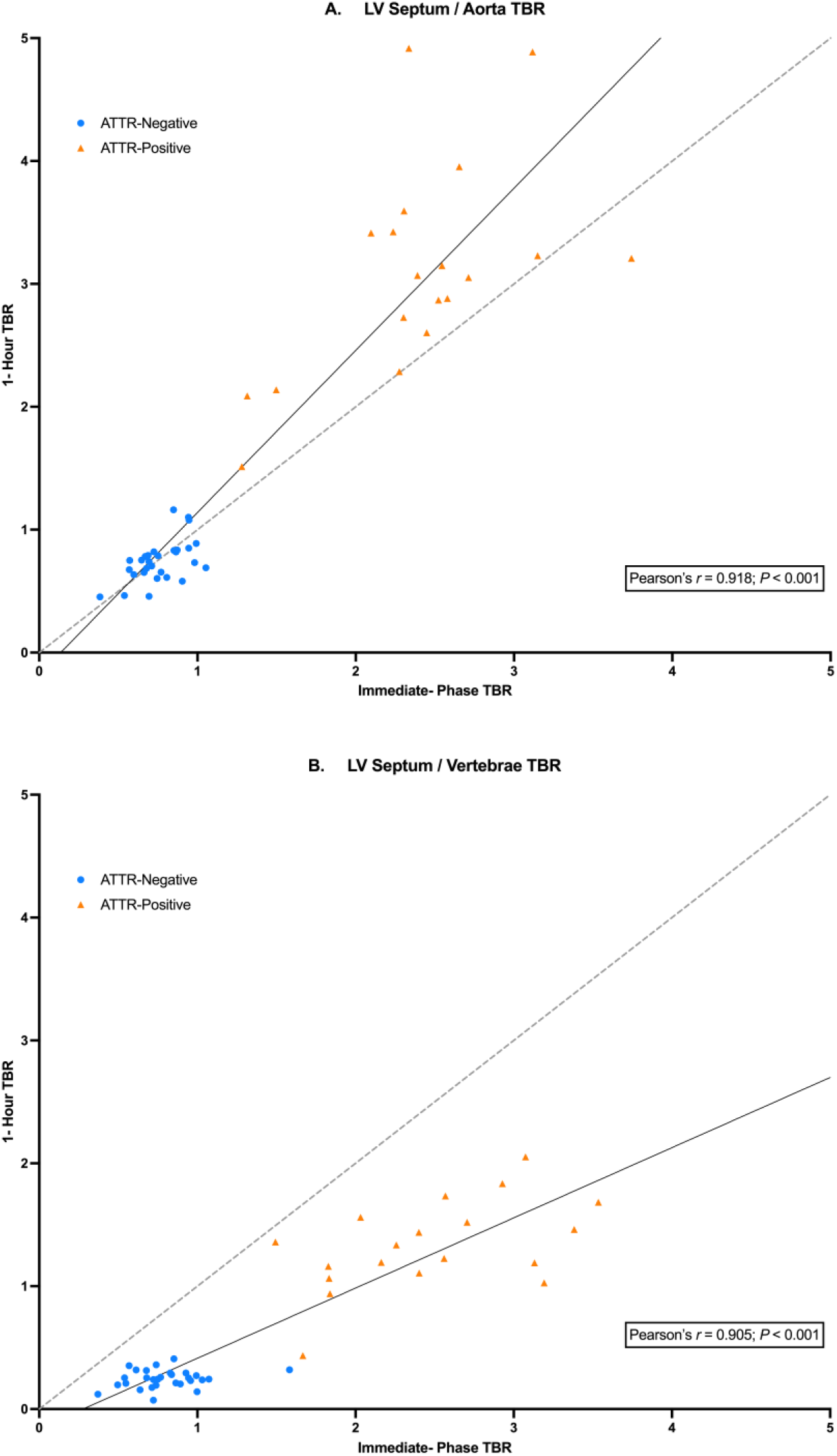
Correlation of ^99m^Tc-HDP SPECT/CT TBRs between immediate and 1-hour acquisitions in A) LV Septum/Aorta TBR and B) LV Septum/Vertebrae TBR **Abbreviations:** ATTR-CM, transthyretin amyloid cardiomyopathy; LV, left ventricular; SPECT/CT, single-photon emission computed tomography/computed tomography; TBR, target-to-background ratio; ^99m^Tc-HDP, technetium-99m hydroxymethylene diphosphonate.

**Figure 3.**
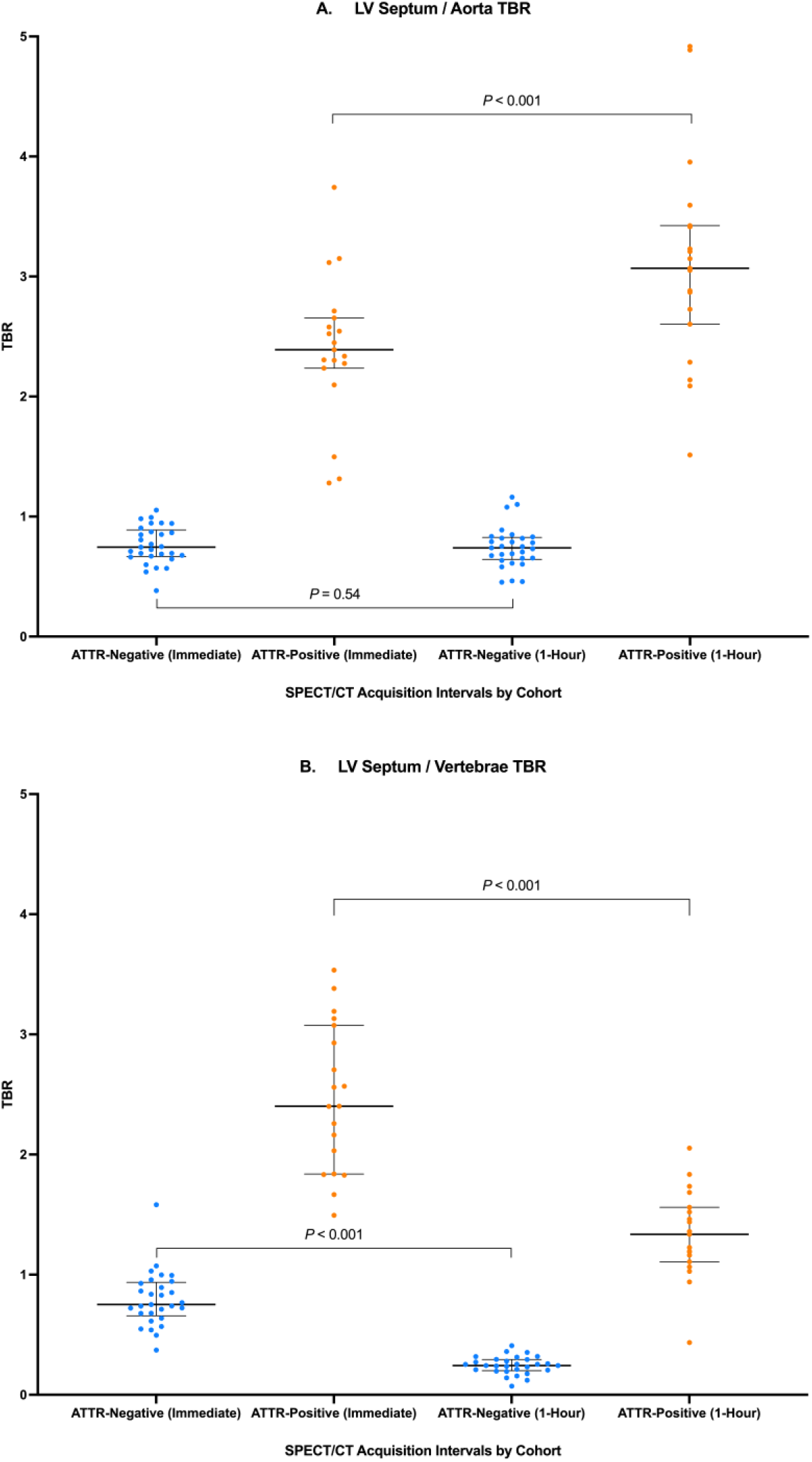
Comparison of TBR changes between immediate and 1-hour ^99m^Tc-HDP SPECT/CT acquisitions in A) LV Septum/Aorta TBR and B) LV Septum/Vertebrae TBR **Abbreviations:** ATTR-CM, transthyretin amyloid cardiomyopathy; LV, left ventricular; SPECT/CT, single-photon emission computed tomography/computed tomography; TBR, target-to-background ratio; ^99m^Tc-HDP, technetium-99m hydroxymethylene diphosphonate.

### Vertebrae Reference

These measures provided the lowest degree of correlation between timepoints, *r* = 0.905 (95% CI: 0.835 to 0.946; *p* < 0.001), Table 2 and Figure 2B, and also differentiated between cohorts during immediate-phase imaging (ATTR-negative: 0.80 ± 0.23 vs ATTR-positive: 2.47 ± 0.61; *p* < 0.001). A significant decrease in mean TBRs for both the ATTR-negative (−0.55; 95% CI: −0.64 to −0.46; *p* < 0.001) and ATTR-positive cohorts (−1.14; 95% CI: −1.39 to −0.89; *p* < 0.001) was observed, Table 3 and Figure 3B. Individual patient TBR data across all anatomical reference regions are provided in Supplemental Tables 1 and 2.

### Inter-observer and Intra-observer Variability

There was excellent agreement (*κ* = 1.000; CI: 1.00 to 1.00; *p* < 0.001) in binary diagnostic classification for both blinded intra-observer and inter-observer agreement across 10% of the total cohort that was randomly sampled. Specifically, no change in diagnostic classification was observed between repeated evaluations by the primary reader or between independent evaluations by two separate readers at either timepoint.

## Discussion

In this prospective evaluation of ^99m^Tc-HDP SPECT/CT, an immediate-phase ^99m^Tc-HDP SPECT/CT acquisition demonstrated similar diagnostic concordance with a standard 1-hour acquisition for the evaluation of suspected ATTR-CM. Our results suggest that an immediate-phase SPECT/CT acquisition may be feasible and warrants further consideration in clinical imaging workflows. This is likely driven by the advantageous pharmacokinetic profile of ^99m^Tc-HDP, where rapid blood-pool clearance occurs due to efficient skeletal redistribution [4,8,9,12], enabling visual contrast that is distinct from residual background activity within minutes of tracer administration. While consensus recommendations historically emphasized multi-hour, delayed acquisition intervals to ensure adequate blood-pool clearance for purposes of visual grading on planar acquisitions, 1-hour ^99m^Tc-PYP scintigraphy was only validated when used in conjunction with semi-quantitative H/CL ratios [3]. Subsequent investigation has since established high diagnostic agreement between one and 3-hour intervals utilizing ^99m^Tc-PYP SPECT [7]. However, our results show that traditional acquisition intervals may not be necessary for diphosphonates like ^99m^Tc-HDP, which possesses distinct pharmacokinetic properties that can be fully leveraged when used in conjunction with SPECT/CT [4,8,9,12].

While previous studies have investigated the favorable kinetics of ^99m^Tc-HDP for accelerated planar imaging [10,11], our data show that immediate-phase planar scintigraphy is inherently limited by poor specificity due to its inability to differentiate true myocardial tracer uptake from residual LV blood-pool activity. However, modern SPECT/CT alleviates this limitation by providing the anatomical localization and the necessary resolution to separate the myocardial wall from the ventricular cavity [13]. This technological advantage allows for the precise quantification of early tracer localization, potentially bypassing the need for extended wait times. The high diagnostic concordance and correlation of our SPECT/CT acquisitions between timepoints builds upon the work of Tersalvi *et al.*, who established that 1-hour ^99m^Tc-HDP SPECT/CT is feasible with excellent diagnostic capabilities [6]. The physiological rationale for immediate ^99m^Tc-HDP imaging is further supported by the recent works of Sperry *et al.*, who demonstrated that diagnostic myocardial uptake of ^99m^Tc-HDP in ATTR-CM occurs within 10 minutes of injection [4,12]; a timeframe that is consistent with a similar investigation that utilized ^99m^Tc-PYP [14]. Furthermore, Sperry’s modern investigation of the immediate-phase kinetics of both ^99m^Tc-HDP and ^99m^Tc-PYP established that while both tracers share similar myocardial retention, ^99m^Tc-HDP is characterized by faster blood-pool clearance and earlier skeletal uptake [4]. Our findings extend these observations and support the clinical feasibility of a streamlined, immediate-phase imaging protocol that could be effectively translated into clinical practice.

While the exact binding mechanism of myocardial-avid tracers in ATTR-CM is unknown, the current study illustrates the dynamic nature of ^99m^Tc-HDP myocardial uptake and tissue redistribution. Our data suggest that the diagnostic hallmark of ATTR-CM, or lack thereof, is established almost immediately after tracer administration. This is evidenced by the high degree of correlation observed between the immediate and 1-hour scans across all reference sites. The tracer redistribution underlying our quantitative TBR measurements is illustrated by the divergent trends observed between cohorts. While the LV Septum/Aorta TBR in the ATTR-positive cohort increased significantly—driven primarily by the rapid clearance of ^99m^Tc-HDP from the vasculature relative to myocardial uptake—the LV Septum/Aorta TBR in the ATTR-negative cohort remained stable, providing a reliable baseline for quantitative assessment. The importance of this stable reference is underscored by the dynamic nature of the LV Septum/Vertebrae TBR, which significantly decreased across both cohorts over time due to the remarkable skeletal avidity of ^99m^Tc-HDP.

Immediate-phase SPECT/CT acquisitions may offer further advantages that extend beyond their potential to improve laboratory throughput and increase patient satisfaction [15]. It is yet to be established which specific timepoint performs optimally in terms of quantification and diagnosis, however, prior studies have demonstrated that planar scintigraphy using visual grading alone may fail to detect cardiac involvement in certain ATTR genetic variants [7,16]. Recent kinetic investigation suggests that prolonged imaging delays can confound visual interpretation of SPECT/CT acquisitions, as progressive skeletal dominance driven by tracer accumulation in the bone occurs significantly earlier with ^99m^Tc-HDP when compared to ^99m^Tc-PYP [4]. Because default image scaling often normalizes to the brightest voxels within a given scan, intense skeletal uptake can artificially suppress visualized myocardial uptake, potentially confounding grading in borderline cases [4]. This visual ambiguity is reflected in the findings of Tersalvi *et al*., where 3-hour ^99m^Tc-HDP SPECT/CT acquisitions required significantly more third-party arbitration compared to 1-hour scans [6], highlighting that even three-dimensional tomography remains vulnerable to visual artifact related to skeletal avidity [4]. As a result of the distinct pharmacokinetics of ^99m^Tc-HDP [4,8,9,12], the immediate-phase imaging window may capture myocardial uptake during the moments of peak visual contrast. As such, further investigation into these immediate-phase dynamics and their correlation with cardiac structure and function is warranted.

As clinician and public awareness of the prevalence of ATTR-CM continues to grow, nuclear laboratories wishing to optimize workflow efficiency can consider evaluating a similar protocol. Accelerated SPECT/CT workflows such as this may offer a practical alternative for the modern evaluation of ATTR-CM.

### Study Limitations

This protocol employed hybrid SPECT/CT. Consequently, our findings may not be generalizable to nuclear laboratories with cardiac and chest SPECT imaging alone. Additionally, we evaluated the diagnostic concordance between imaging timepoints rather than establishing *de novo* diagnoses of ATTR-CM. As such, the absolute diagnostic accuracy of immediate-phase imaging against a gold standard diagnosis—whether pathological or by established non-invasive criteria—remains to be determined in future studies. Larger studies are required to confirm our findings, especially in cohorts of early disease, given the evolving understanding of sensitivity and specificity in this specific population.

## Conclusion

In this cohort of patients with suspected ATTR-CM, immediate ^99m^Tc-HDP SPECT/CT demonstrated high diagnostic concordance with standard 1-hour protocols by qualitative and quantitative metrics. This expedited approach may reduce scan times, improve nuclear laboratory throughput, and increase patient satisfaction.

## Data Availability

All data produced in the present study are available upon reasonable request to the authors.

---

**Supplemental Figure 1.**
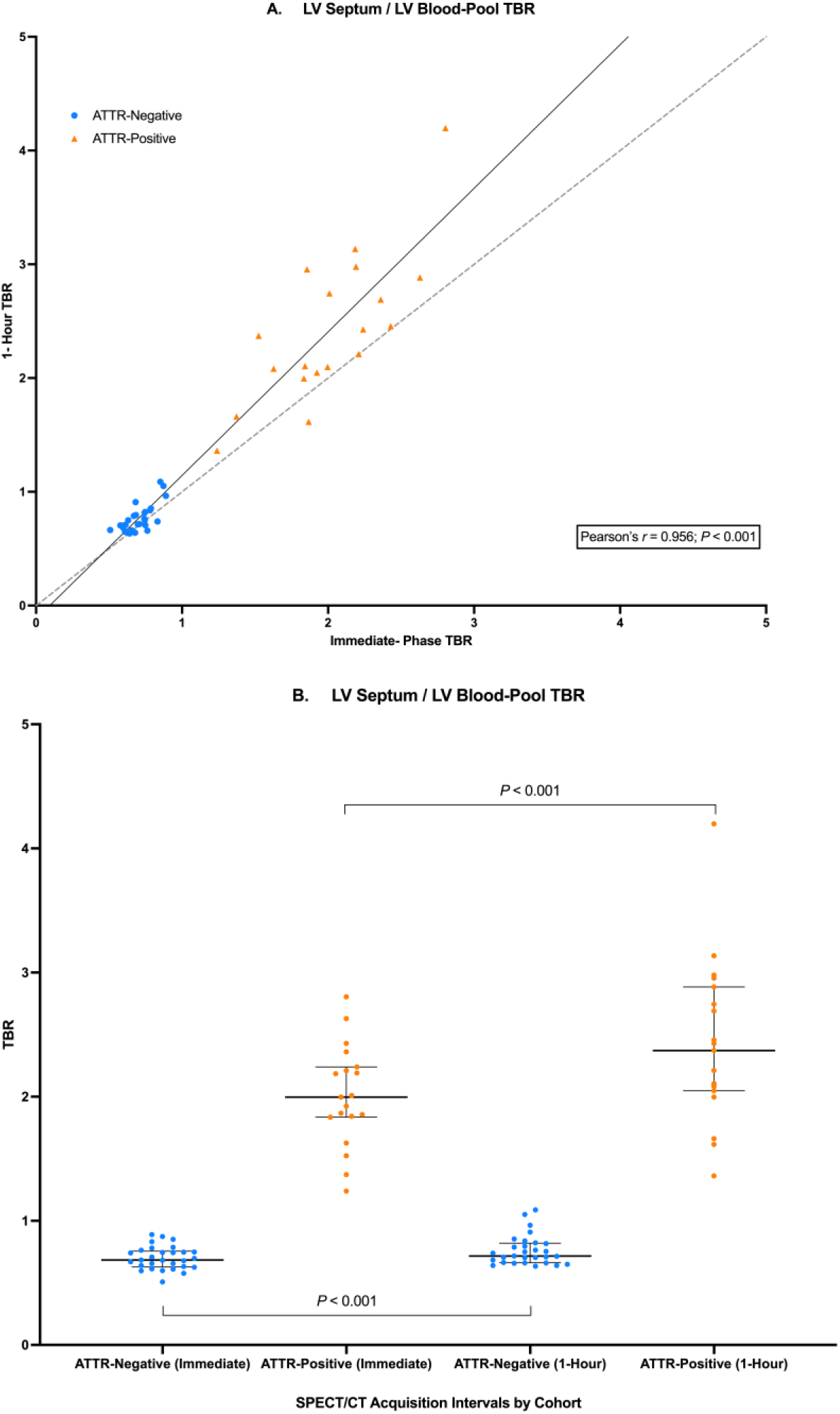
LV Septum/LV Blood-Pool TBR analysis A) correlation of TBRs between immediate and 1-hour ^99m^Tc-HDP SPECT/CT acquisitions and B) Comparison of TBR changes between immediate and 1-hour ^99m^Tc-HDP SPECT/CT acquisitions **Abbreviations:** ATTR-CM, transthyretin amyloid cardiomyopathy; LV, left ventricular; SPECT/CT, single-photon emission computed tomography/computed tomography; TBR, target-to-background ratio; ^99m^Tc-HDP, technetium-99m hydroxymethylene diphosphonate.

## Planar Supplement

### Immediate Planar Imaging and Quantitative Analysis

Immediate-phase planar imaging demonstrated high sensitivity of 94.7% (95% CI: 75.4% to 99.1%) but poor specificity of 31.0% (95% CI: 17.3% to 49.2%) in comparison to the 1-hour SPECT/CT reference standard. Quantitative analysis revealed a decrease in mean H/CL ratios across timepoints (immediate: 1.79 ± 0.36 vs 1-hour: 1.51 ± 0.50, *p* < 0.001) with a mean difference of −0.28 (95% CI: −0.41 to −0.16). There was a moderate linear relationship between immediate and 1-hour H/CL ratios (*r* = 0.538; 95% CI: 0.300 to 0.713; *p* < 0.001). Diagnostic concordance between immediate planar and 1-hour SPECT/CT was poor (*κ* = 0.220; 95% CI: −0.03 to 0.47; *p* = 0.09).

**Supplemental Table 1.**
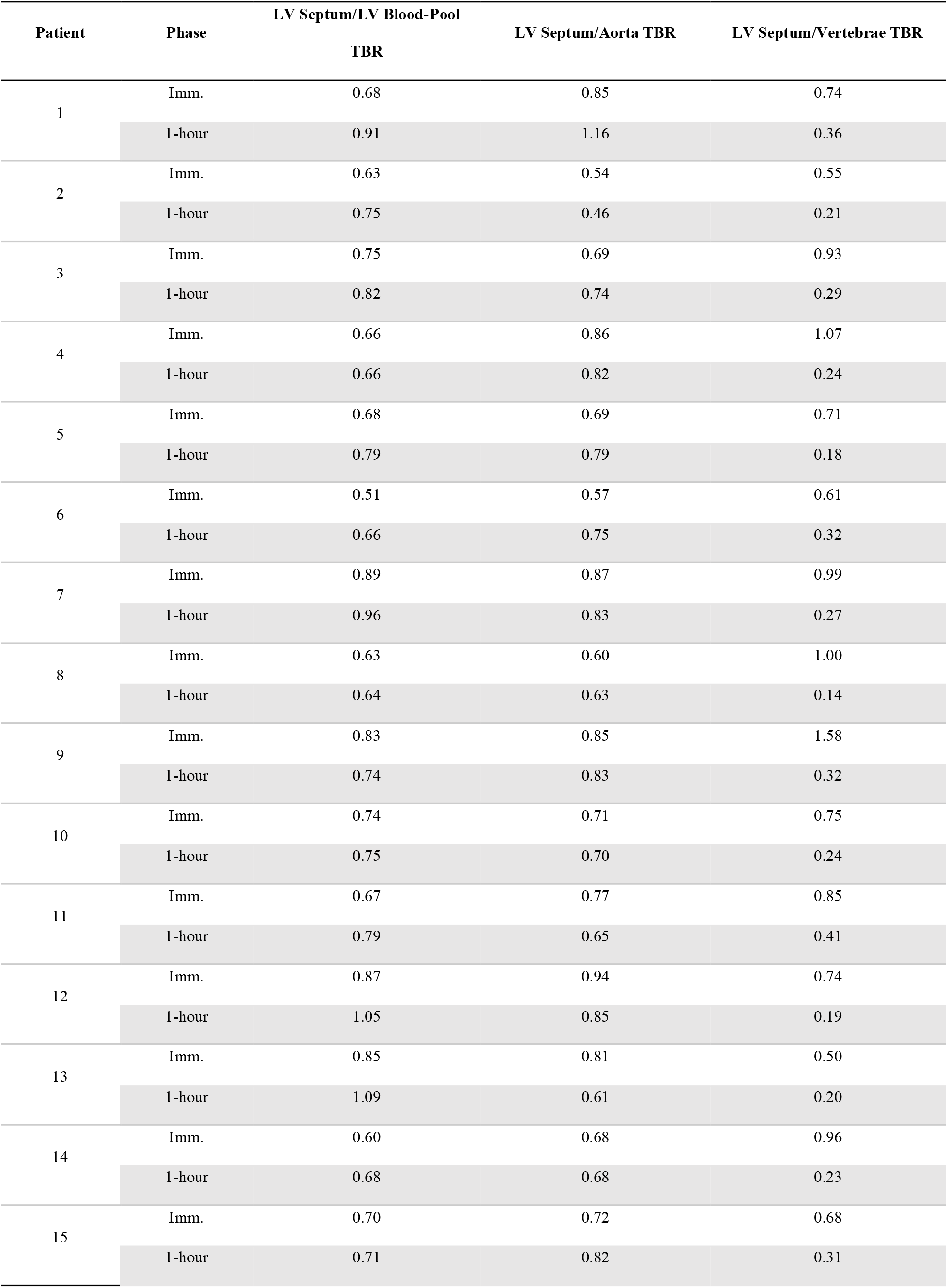
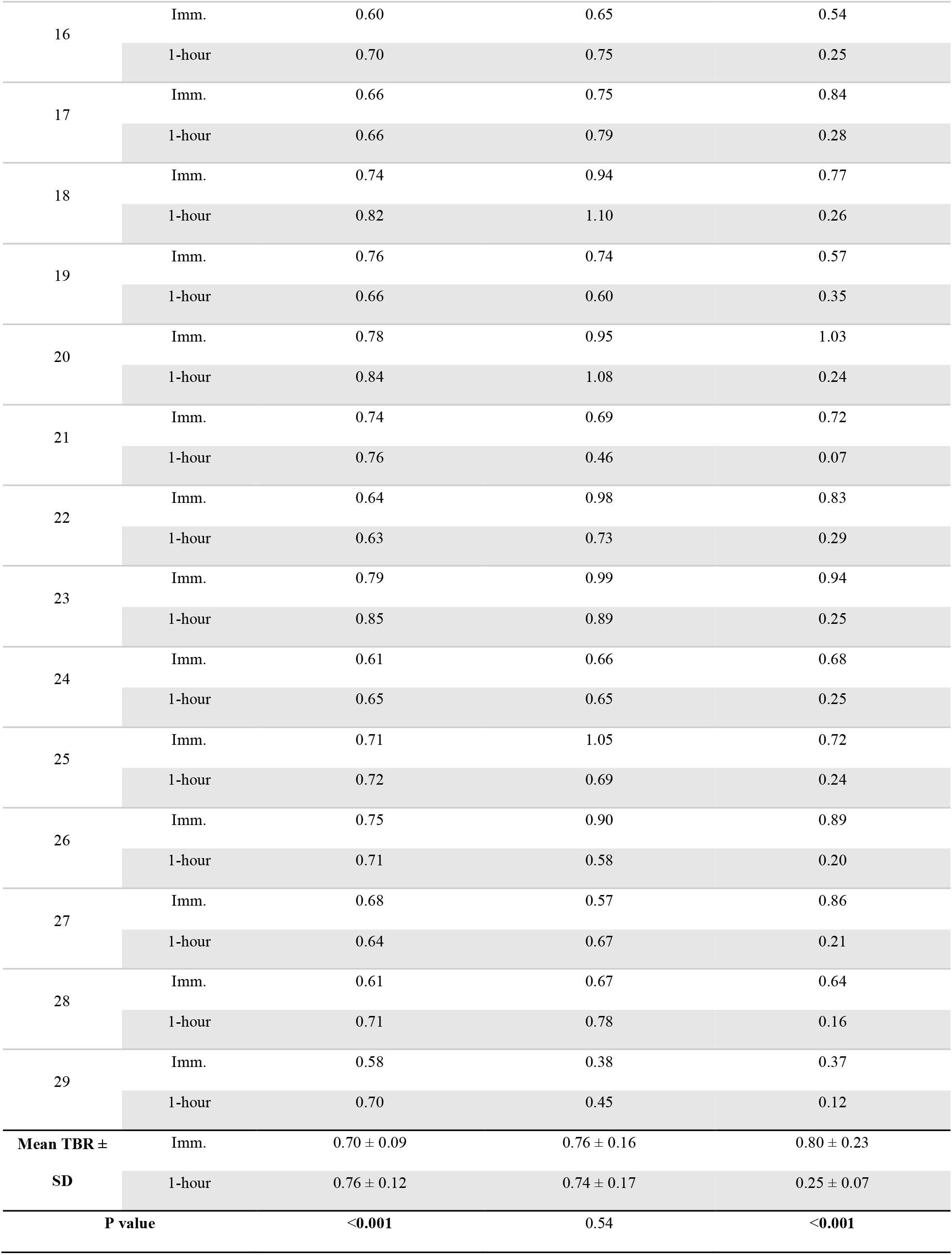
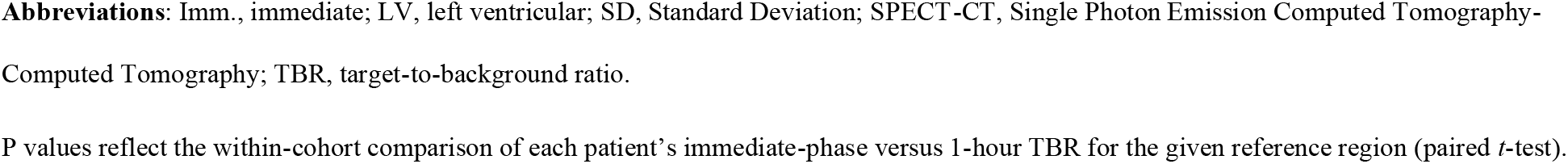
Comprehensive 99mTc-HDP SPECT/CT TBR data for the ATTR-negative cohort.

**Supplemental Table 2.**
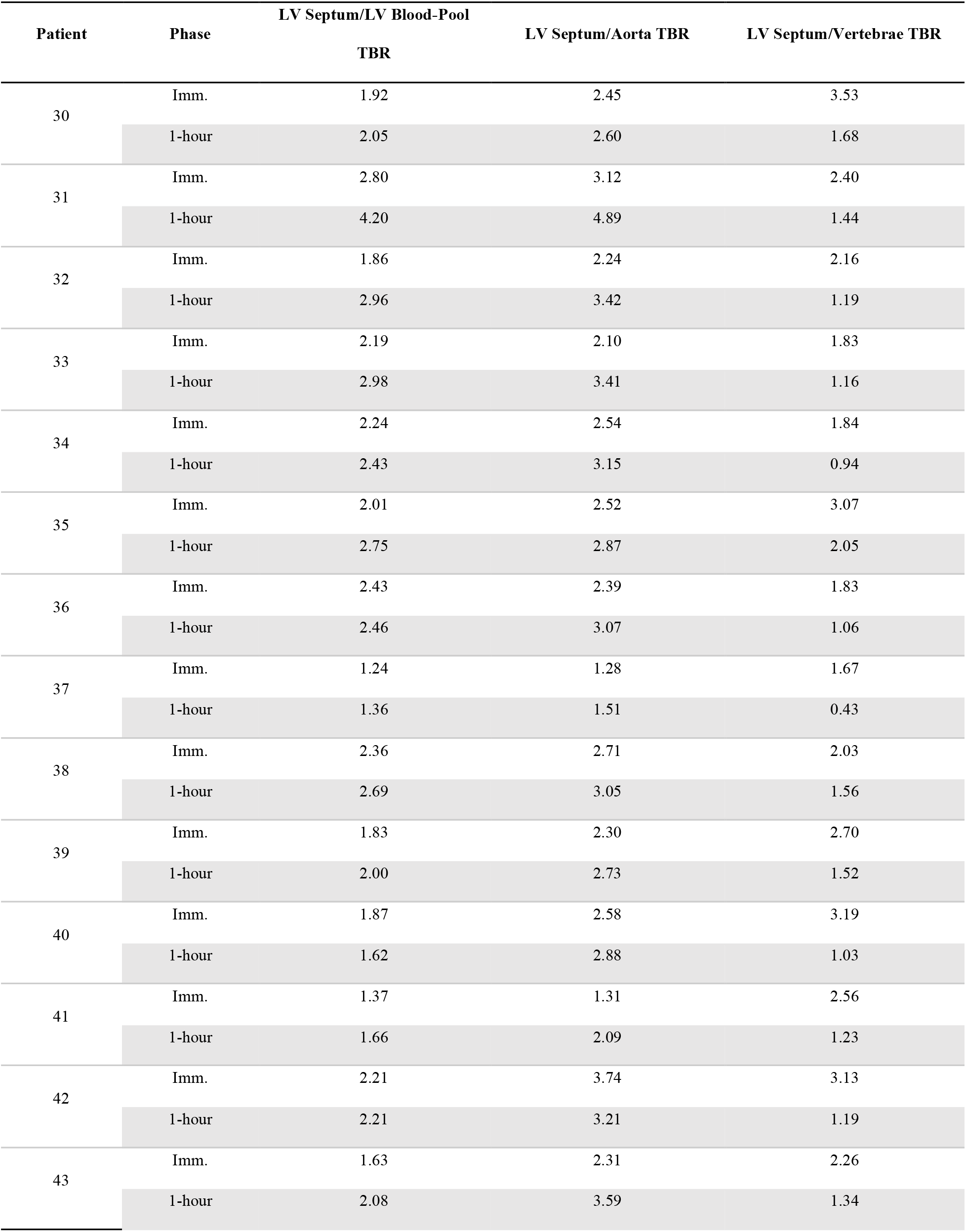
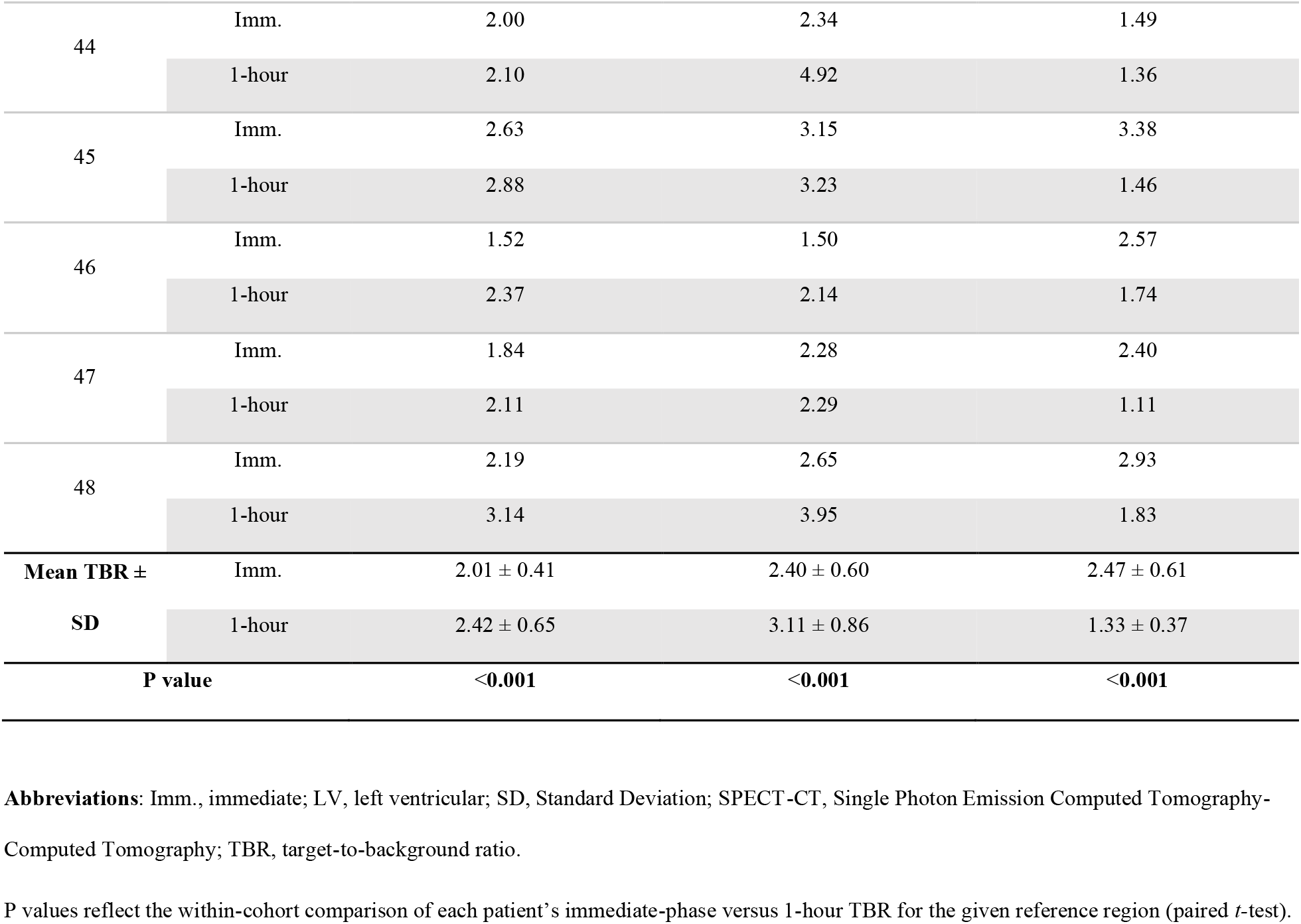
Comprehensive 99mTc-HDP SPECT/CT TBR data for the ATTR-positive cohort.

